# Household energy poverty and sleep disorders among people living with HIV: Does the level of physical activity have any moderating effect?

**DOI:** 10.64898/2026.07.29.26359226

**Authors:** Hefot Ahmed, Mavis Odei Boateng, Eyram Adzo Agbe, Patrick Mbullo Owuor, Ellis Adjei Adams, Godfred Odei Boateng

## Abstract

**Objectives:** Household energy poverty has been associated with deleterious health outcomes, including poor sleep outcomes. However, no study has examined this relationship among people living with HIV (PLHIV) and the possible moderating effect of physical activity. Therefore, this study investigated the relationship between household energy poverty and sleep disorders and assessed whether various levels of physical activity (vigorous, moderate, and minimal) would moderate this relationship among PLHIV in Kenya.

**Methods:** Data for this study were drawn from the Resource Insecurity and Well-being Study on PLHIV in Kenya (N = 1,132), which included information on Insomnia, household energy poverty, a measure of physical activity levels, and demographic measures. Following univariate and bivariate analysis, we conducted multivariate linear regressions followed by interaction models with each level of physical activity.

**Results:** Of the 1132 participants, 74.2% had energy poverty scores above the mean (18.15), and 36.6% reported Insomnia scores of 7.03 and above. In the multivariate model, household energy poverty was associated with greater insomnia scores (β = 0.10, 95% CI: 0.07, 0.12). Vigorous, moderate, and minimal physical activity were disparately associated with lower insomnia scores. Interactions between household energy poverty and physical activity were not statistically significant, although the model suggested a buffering effect.

**Conclusion:** Household energy poverty increases Insomnia, and engaging in any level of physical activity reduces Insomnia among PLHIV. Physical activity has the potential to buffer the effects of energy poverty on Insomnia. These findings suggest that public health interventions should address both structural and lifestyle factors to improve sleep outcomes in vulnerable populations.

## Introduction

Sleep is a vital component of an individual’s life (1). Sleep is essential for maintaining optimal physical, mental, and emotional health, as well as overall well-being (3,4). To maintain normal social functioning, an individual must obtain optimal sleep (6). According to the American Academy of Sleep Medicine, the Sleep Research Society, and the National Sleep Foundation, adults are recommended to get 7-9 hours of sleep every night to maintain optimal health (7,8). The Sleep Foundation considers good quality sleep to be falling asleep within 30 minutes, sleeping through the night with no more than one waking episode, and feeling rested and rejuvenated upon waking in the morning (9).

Optimal sleep is a foundational physiological process for human survival (10), and a lack of optimal sleep can increase an individual’s risk of developing a wide range of diseases and reduce their cognitive functioning (11). However, optimal sleep is difficult to achieve for many people around the world, with 40% of the global population experiencing poor sleep quality and 29% experiencing Insomnia (12). This could be even more severe for people living with HIV (PLHIV) (13). HIV can disrupt sleep through direct and indirect pathways. The direct pathway is driven by the immune system’s response to the virus. In contrast, the indirect pathway is caused by the neurotoxic effects resulting from the immunopathological response in the central nervous system (CNS) (14). Antiretroviral medications, such as Efavirenz, can cause sleep disruptions, including Insomnia (15).

Several factors negatively impact sleep, which can be grouped into environmental, biological, and social factors (16). Environmental factors such as cold conditions (17), transportation noise due to outside traffic (18), and increasing levels of Carbon Dioxide, PM2.5, and temperature have been associated with poor sleep (19). Biological factors, such as disruptions in circadian rhythms, can lead to poor sleep quality (20), which in turn can contribute to metabolic dysregulation and hormonal imbalances (21). Next, social factors such as lack of social cohesion and feeling unsafe in one’s neighbourhood (22), stress (23), and living in poorer neighbourhoods increase the odds of poor sleep outcomes (24). While these social determinants are significant, material circumstances, such as energy insecurity-related issues, may present a significant risk to adverse sleep outcomes.

A plethora of research has examined the determinants of sleep disorders (25–27); however, to the best of our knowledge, no study has examined the effect of household energy poverty on sleep disorders among PLHIV. Energy poverty, a sub-component of energy insecurity, refers to the inability of households to access sufficient and affordable energy sources to meet their basic needs (30). Energy poverty significantly impacts millions of people world (29). Household energy poverty is a growing public health threat and is associated with deleterious health outcomes (30). This growing burden is especially prominent in sub-Saharan Africa (31), where about 970 million Africans lack access to clean cooking fuels (32) and 600 million lack access to electricity (33), with the electricity access rates in 23 African countries below 50% (34). Persistent public power supply interruptions have led many residents to rely on alternative power sources, such as backup diesel generators, to address electricity shortages (35). However, diesel generators can cause noise pollution, which can disrupt sleep by triggering a stress response (36,37). In addition to this, households experiencing energy poverty often adopt risky coping strategies.

These risky coping strategies not only pose significant health risks but also contribute to stress and anxiety, particularly in sub-Saharan Africa, which can negatively impact sleep quality (38,39). Examples of common coping strategies include using space heaters or ovens for warmth (40) (41), reducing spending on food and medicine (42), and keeping homes at unhealthy temperatures (43). Stress, anxiety, and consequently, poor sleep quality, may be heightened by the financial strain of high energy costs, trade-offs between spending money on energy or other necessities, and participating in unsafe heating practices (44). Furthermore, the reliance on solid fuels such as wood for cooking creates indoor air pollution (45), which makes households inconducive for sleep (46,47). Low-income households also struggle to afford air conditioning, which leaves rooms very warm or overheated during hot temperatures (48). In colder regions, the lack of access to energy means people must sleep in extremely cold conditions, which also negatively impacts their sleep. These factors make sleep difficult and could lead to chronic sleep disorders over time (49).

This notwithstanding, engagement in physical activity has been found to improve sleep quality (50) by increasing the production of melatonin, reducing stress and anxiety, and regulating body temperature (51,52). The production of melatonin facilitates better sleep outcomes through three ways: (a) its chronobiotic effect on the suprachiasmatic nucleus (SCN), (b) regulation of the circadian rhythm and sleep-wake cycle, and (c) signalling to tissues and organs that it is nighttime (53). Again, physical activity aids in regulating body temperature, a key factor in promoting sleep onset. Sleep is more easily initiated when core body temperature declines, while it is difficult to maintain sleep when the core body temperature is rising (54). While these physiological processes highlight how physical activity can play a key role in improving sleep quality, it is essential to examine how various levels of physical activity influence sleep outcomes.

Regular engagement in vigorous and moderate physical activity is associated with a reduced risk of developing metabolic syndrome (55). This condition is often accompanied by sleep abnormalities (56). Also, light physical activity in the daytime has been associated with improved sleep quality (57) and a significant reduction in postprandial glucose (58), which is associated with better sleep quality (59), Based on this understanding, this study aims to 1) examine the relationship between household energy poverty and sleep disorders; 2) the relationship between the various levels of physical activity and sleep disorders, and 3) the moderating effect of the different levels of physical activity on sleep disorder among PLHIV in Kenya.

## Methods

### Ethics Statement

We received IRB approval from the Institutional Review Boards at the University of Texas at Arlington and the University of Notre Dame (21-05-6613). Also, ethical approval was obtained from the Africa Medical Research Institute (AMREF) Ethics and Scientific Research Committee (ESRC-P1396-2023). Data collection on measures used in this study started July 10, 2023, and ended August 3, 2023. All participants provided verbal informed consent that was documented as part of the data collection process via the Computer-Assisted Personal Interview. Agreements to engage in the research were recorded as consent as part of data collection. This approach to obtaining consent was approved by all the Ethics Committees. Minors were not included in this study. We confirm that the study complied with ethical standards outlined in the Belmont Report and/or Declaration of Helsinki

### Study setting and population

This study was conducted in collaboration between the Pamoja Community-Based Organization (CBO) in Kenya, the University of Texas at Arlington, and the University of Notre Dame. Pamoja CBO is a grassroots organization with the primary role of supporting communities in addressing the needs that impact their health and well-being (60). The Resource Insecurity and Well-being study, a collaborative project with Pamoja CBO, was conducted in the Kisumu West and Seme sub-counties of Kisumu County, Kenya, where Pamoja had been implementing HIV prevention interventions for the past decade (60). This study employed a cross-sectional survey design to examine the self-reported physical activity behaviours of PLHIV. The analytical sample size varied, ranging from 1,051 to 1,132 participants, due to differences in response rates across variables and data cleaning.

### Study design and data collection

The purpose of this study was to examine the direct and indirect relationship between household energy poverty and sleep disorders among people living with HIV in Kenya, while accounting for the possible moderating role of physical activity. Before participating in the study, all participants gave verbal informed consent. Between July 10, 2023, and August 3, 2023, data were collected from Seme sub-counties (n = 766) and Kisumu West sub-counties (n = 366), totalling a sample size of 1,132 PLHIV. The survey was administered using a stratified two-stage cluster sampling method, and households were randomly selected based on a manual mapping of the areas where the CBO served PLHIV. Surveys and interviews were limited to participants aged 16 years and older. Data were collected on participants’ sociodemographic characteristics, environmental risk factors, resource insecurity, sleep health, and mental health conditions.

### Outcome measures

The outcome variable for this study was sleep disorder, specifically Insomnia, which was measured using the first five questions of the sleep disorder questionnaire. These questions asked participants whether they had trouble falling asleep, staying asleep, needed to take anything to help them sleep, used alcohol to help them sleep, and the frequency with which any medical conditions disrupted their sleep. With Likert-type responses ranging from 1 (never) to 5 (always), we created a score for Insomnia, which ranged from 5 to 17, and a Cronbach’s alpha of 0.75, exceeding the recommended threshold of 0.70. Thus, higher scores on this scale indicate an increasing prevalence of sleep disorders.

### Moderators

Vigorous Physical Activity. We assessed various levels of physical activity for our moderator – vigorous, moderate, and minimal – using the International Physical Activity Questionnaire (IPAQ) Short-Form (61). This measured physical activity in terms of vigorous, moderate, walking, and sitting over the past 7 days. Frequency (days per week) and duration (hours per day) were assessed for each type of physical activity, except for sitting time, which was measured in terms of hours spent sitting on weekdays (61). For this vigorous activity, participants were asked, “During the last 7 days, on how many days did you do vigorous activities like heavy lifting, digging, aerobics, or fast bicycling (61)? How much time did you spend doing vigorous activities on one of those days (hours per day) (61)?” To create the score, we multiplied the number of days by the time spent doing those activities. Higher scores suggest greater vigorous physical activity, while lower scores suggest lower vigorous physical activity.

Moderate Physical Activity. For this measure, participants were asked, “During the last 7 days, on how many days did you do moderate physical activities like carrying light loads, bicycling at a regular pace, or doubles tennis (61)? How much time did you usually spend doing moderate physical activities on one of those days (61)?” The number of days was multiplied by the amount of time spent doing those activities to generate a score.

Minimal Physical Activity. Participants were asked about walking activity “during the last 7 days, on how many days did you walk for at least 10 minutes at a time (61)? How much time did you usually spend walking on one of those days (61)?” A score for minimal physical activity was created by multiplying the number of days by the amount of time spent engaging in those activities.

### Explanatory variable

We used the Household Energy Poverty Experiences Scale, a 10-item scale validated by Boateng and Armah (81). Among the battery of questions, participants were asked whether they worried that their energy will be used up before they got money to refill, the frequency with which they lacked money to buy energy, whether they reduced energy consumption to uncomfortable levels because there was no money to buy energy, reduced expenses because there was not enough money to buy basic household needs and pay for energy, and whether they had to change what was been eaten because there were problems with energy. This scale produced a score ranging from 0 to 30, with a Cronbach’s alpha of 0.93. Higher scores suggest higher energy poverty.

### Control variables

Socio-cultural and biosocial factors were considered as covariates to understand better the factors that could influence the relationship between energy poverty and sleep disorders. Socio-cultural factors included the number of children under 5 years old living in the house. Marital status was categorized as single or unmarried (1), separated/divorced/widowed (2), and married or living with a partner (3). Place of residence (neighbourhood) was coded as (1) Seme sub-county and (2) Kisumu sub-county. Level of education was categorized as (0) no education, (1) primary school, (2) middle or junior high school, (3) secondary or high school, and (4) college or university. Biosocial factors included the participants’ age and gender. Age was used as a count variable, while gender was coded as male (0) and female (1).

### Data Analysis

Data analysis incorporated descriptive, bivariate, multivariate, and moderation techniques. We described the distribution of our study variables using percentages and means with standard deviation. We then examined the bivariate relationship between predictor variables and sleep disorders using a linear regression model. Using multivariate linear regression, we examined the relationship between household energy poverty and sleep disorders, controlling for covariates such as biosocial and socio-cultural factors. We then assessed the moderating effect of various levels of physical activity on the relationship between household energy poverty and sleep disorders. We examined the relationship between household energy poverty and each level of physical activity to determine whether it had a buffering effect on sleep disorders, while adjusting for covariates. STATA version 19 (Stata Corp., College Station, Texas, USA) was used for all data analysis.

## Results

### Sample characteristics

Table 1 shows the general distribution of the variables. The study sample consisted of 1,132 people living with HIV in Kenya. The mean household energy poverty score was 18.15 (SD = 6.66). In terms of gender distribution, a large portion were female (88.07%), while only 11.93% were male. Nearly half of the participants were separated, divorced, or widowed (48.85%), and a small portion were single (5.39%). The majority resided in the Seme Sub-Counties (67.67%), while 32.33% resided in the Kisumu West Sub-Counties. Most participants completed primary education (73.59%), followed by middle/junior high school (19.17%), secondary/senior high school (1.50%), and tertiary education (1.33%). The mean socioeconomic status score was 3.01 (SD = 1.27), and the average monthly Income was 5,870.53 (in KES) on a scale of 0-60,000. The mean score for Insomnia was 7.03 (SD = 2.60). Participants engaged in a mean of 996.57 minutes per week of vigorous physical activity (SD = 877.40).

**Table 1.** Sample Characteristics (N = 1,132)

| Descriptives | Mean (SD)/ n (%) |
| --- | --- |
| **Key Predictors** |  |
| Household Energy Poverty | 18.15 (6.66) |
| **Biosocial Factors** |  |
| Age | 44.04 (12.45) |
| *Gender* |  |
| Male | 135 (11.93) |
| Female | 997 (88.07) |
| Number of Children (<5years) | 0.73 (1.46) |
| **Socio-Cultural Factors** |  |
| *Marital Status* |  |
| Single | 61(5.39) |
| Separated/divorced/widowed | 553(48.85) |
| Married/cohabiting | 518 (45.76) |
| *Neighborhood* |  |
| Seme Sub-Counties | 766 (67.67) |
| Kisumu West Sub-County | 366 (32.33) |
| *Education* |  |
| No Education | 50 (4.42) |
| Primary | 833 (73.59) |
| Middle/Junior High | 217 (19.17) |
| Secondary/Senior High | 17 (1.50) |
| Tertiary | 15 (1.33) |
| Subjective Socioeconomic Status | 3.01 (1.27) |
| Average Income (monthly) - x (range) | 5870.53 (0-60000) |
| Health Status (Poor Health) | 1.62 (0.67) |
| **Outcome variables** |  |
| Insomnia | 7.03 (2.60) |
| **Moderators** |  |
| Vigorous Physical Activity | 996.57 (877.40) |
| Moderate Physical Activity | 441.90 (621.07) |
| Minimal Physical Activity | 543.13 (670.06) |

### Bivariate regression between predictor variables and Insomnia

Table 2 displays the results from bivariate regressions, which found that household energy poverty was linked with higher insomnia scores (β = 0.10, 95% CI: 0.08, 0.13). All levels of physical activity were associated with lower insomnia scores, particularly vigorous physical activity (β = -0.0002456, 95% CI: -0.0004277, -.0000634). Older participants experienced higher levels of Insomnia (β = 0.03, 95% CI: 0.02, 0.04), while females reported lower levels of Insomnia (β = -0.38, 95% CI: -0.89, 0.14). Being married (β = 1.43, 95% CI: 0.97, 1.89) or separated (β = 1.43, 95 % CI: 0.97, 1.89) was associated with higher insomnia scores. Living in Kisumu West was associated with lower insomnia scores (β = -0.75, 95% CI: -1.05 to -0.44). Those with higher socioeconomic status (β = -0.32, 95% CI: - 0.43, -0.20) and Income (β = -0.0000883, 95% CI: -0.00011, -0.0000644) experienced lower insomnia scores respectively, while those with poor health experienced higher levels of Insomnia (β = 0.88, 95% CI: 0.66 to 1.11).

**Table 2.**
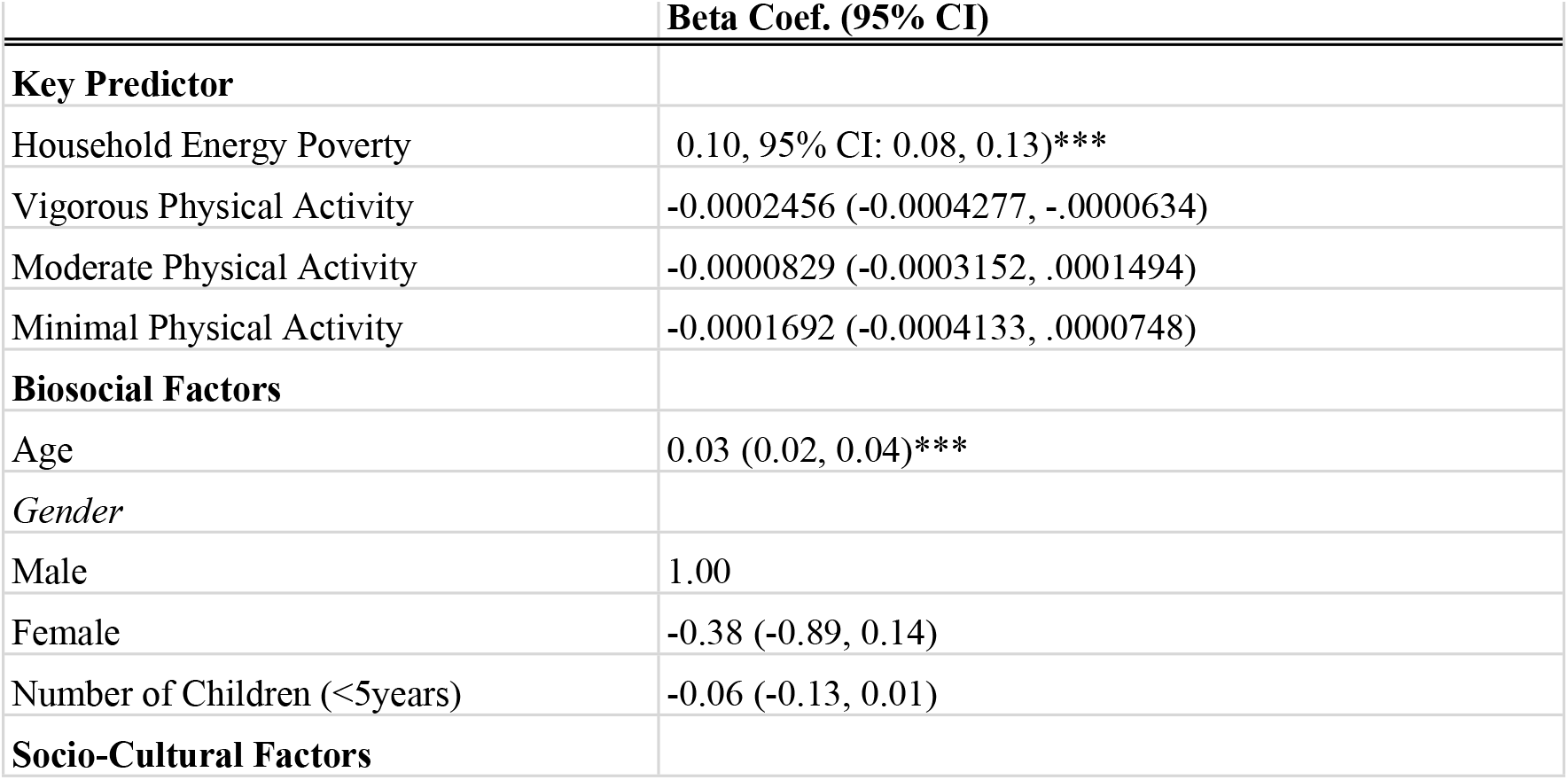
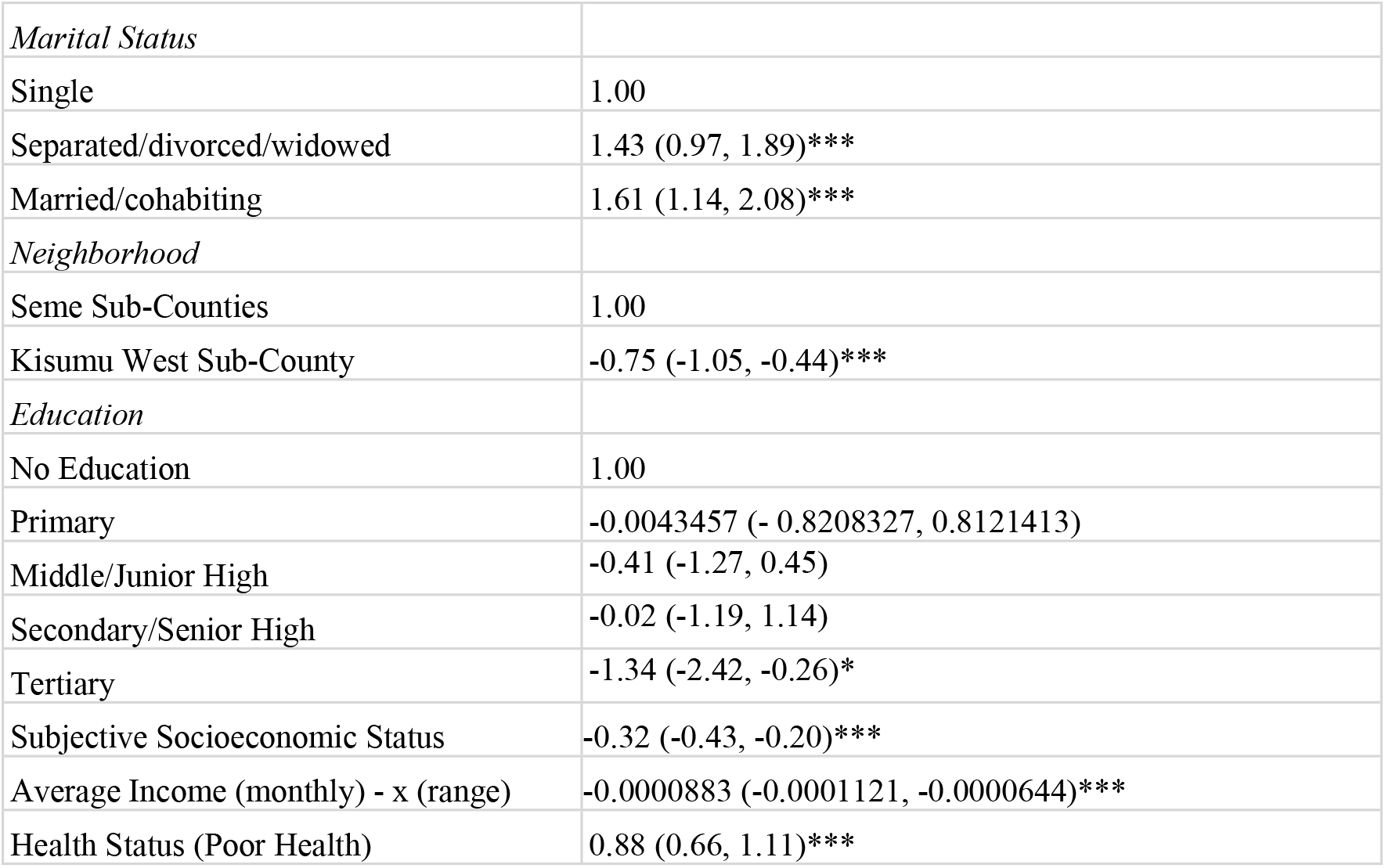
Bivariate regression results between the predictor variables and Insomnia.

|  | Beta Coef. (95% CI) |
| --- | --- |
| **Key Predictor** |  |
| Household Energy Poverty | 0.10, 95% CI: 0.08, 0.13)*** |
| Vigorous Physical Activity | -0.0002456 (-0.0004277, -.0000634) |
| Moderate Physical Activity | -0.0000829 (-0.0003152, .0001494) |
| Minimal Physical Activity | -0.0001692 (-0.0004133, .0000748) |
| **Biosocial Factors** |  |
| Age | 0.03 (0.02, 0.04)*** |
| *Gender* |  |
| Male | 1.00 |
| Female | -0.38 (-0.89, 0.14) |
| Number of Children (<5years) | -0.06 (-0.13, 0.01) |
| **Socio-Cultural Factors** |  |
| *Marital Status* |  |
| Single | 1.00 |
| Separated/divorced/widowed | 1.43 (0.97, 1.89)*** |
| Married/cohabiting | 1.61 (1.14, 2.08)*** |
| *Neighborhood* |  |
| Seme Sub-Counties | 1.00 |
| Kisumu West Sub-County | -0.75 (-1.05, -0.44)*** |
| *Education* |  |
| No Education | 1.00 |
| Primary | -0.0043457 (- 0.8208327, 0.8121413) |
| Middle/Junior High | -0.41 (-1.27, 0.45) |
| Secondary/Senior High | -0.02 (-1.19, 1.14) |
| Tertiary | -1.34 (-2.42, -0.26)* |
| Subjective Socioeconomic Status | -0.32 (-0.43, -0.20)*** |
| Average Income (monthly) - x (range) | -0.0000883 (-0.0001121, -0.0000644)*** |
| Health Status (Poor Health) | 0.88 (0.66, 1.11)*** |

### Multivariate regression between predictor variables and outcome measure

Table 3 presents results from multivariate regression models indicating that as household energy poverty increased, participants experienced higher levels of Insomnia (β = 0.09, 95% CI: 0.07, 0.11). Age was associated with higher levels of Insomnia (β = 0.01, 95% CI: -0.0023, 0.03), while being female was associated with lower levels of Insomnia (β = -0.44, 95% CI: -0.93, 0.06), although this difference was not statistically significant. Similarly, an increase in the number of children aged 5 years and below was associated with a decrease in Insomnia (β = -0.05, 95% CI: -0.21, 0.11). This relationship was not significant. Those who were separated/divorced/widowed experienced higher levels of Insomnia (β = 1.10, 95% CI: 0.40, 1.80). Similarly, individuals who were married/cohabiting experienced higher levels of Insomnia (β = 1.25, 95% CI: 0.61, 1.89), while those living in Kisumu West Sub-Counties experienced lower levels of Insomnia (β = -0.35, 95% CI: -0.67, -0.0301).

**Table 3.** Multivariate linear regression of the relationship between household energy poverty and Insomnia, adjusting for covariates among PLHIV in Kenya (n=1,005)

|  | Beta Coeff. (95% CI) |
| --- | --- |
| **Key Predictor** |  |
| Household Energy Poverty | 0.09 (0.07, 0.11)*** |
| **Biosocial Factors** |  |
| Age | 0.01 (-0.02, 0.03) |
| *Gender* |  |
| Male | 1.00 |
| Female | -0.44 (-0.93, 0.06)* |
| Number of Children (<5years) | -0.05 (-0.21, 0.11) |
| **Socio-Cultural Factors** |  |
| *Marital Status* |  |
| Single | 1.00 |
| Separated/divorced/widowed | 1.10 (0.40, 1.80)** |
| Married/cohabiting | 1.25 (0.61, 1.89)*** |
| *Neighborhood* |  |
| Seme Sub-Counties | 1.00 |
| Kisumu West Sub-County | -0.35 (-0.67, -0.03)* |
| *Education* |  |
| No Education | 1.00 |
| Primary | -0.27 (-1.13, 0.58) |
| Middle/Junior High | -0.31 (-1.25, 0.64) |
| Secondary/Senior High | 0.33 (-0.90, 1.55) |
| Tertiary | -0.37 (-1.62, 0.88) |
| Subjective Socioeconomic Status | -0.25 (-0.38, -0.12)*** |
| Average Income (monthly) - x (range) | -0.0000969 (-0.0001197, -0.0000741)*** |
| Health Status (Poor Health) | 0.73 (0.50, 0.95)*** |
| R<sup>2</sup> | 0.227 |
| Adj R<sup>2</sup> | 0.216 |

The level of education was not significantly associated with Insomnia, except for tertiary education, whose statistical significance was attenuated in the multivariate model. Those who reported higher subjective socioeconomic status (β = -0.25, 95% CI: -0.38, -0.12) and average income (monthly) (β = -0.0000969, 95% CI: -0.0001197, -0.0000741) were associated with lower levels of Insomnia. Those reporting poorer health were associated with higher insomnia scores (β = 0.73, 95% CI: 0.50, 0.95).

Table 4 presents multivariate results that show the various levels of physical activity and their association with Insomnia assessed in separate models. Vigorous physical activity was associated with lower insomnia scores (β = - 0.00041, 95% CI: -0.00063, -0.00019), while higher household energy poverty was associated with greater insomnia scores (β = 0.10, 95% CI: 0.07, 0.12). Moderate physical activity was associated with lower insomnia scores (β = - 0.00040, 95% CI: -0.00065, -0.00015), while household energy poverty was associated with higher insomnia scores (β = 0.10, 95% CI: 0.07, 0.12). Minimal physical activity was associated with lower insomnia scores (β = -0.00025, 95% CI: -0.00050, -0.00000564), while household energy poverty was positively associated with Insomnia (β = 0.09, 95% CI: 0.07, 0.12).

**Figure 1.**
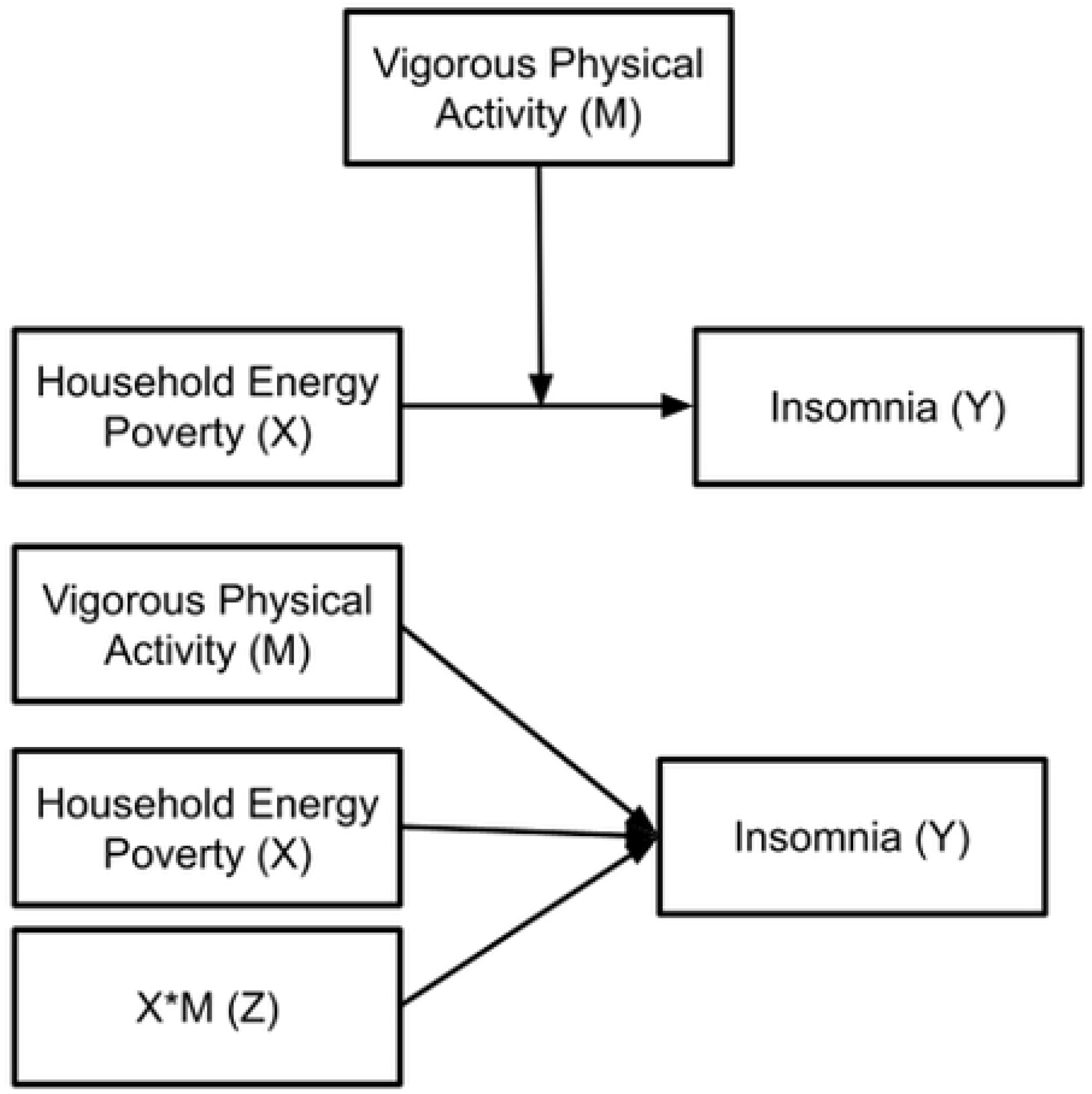

**Table 4.**
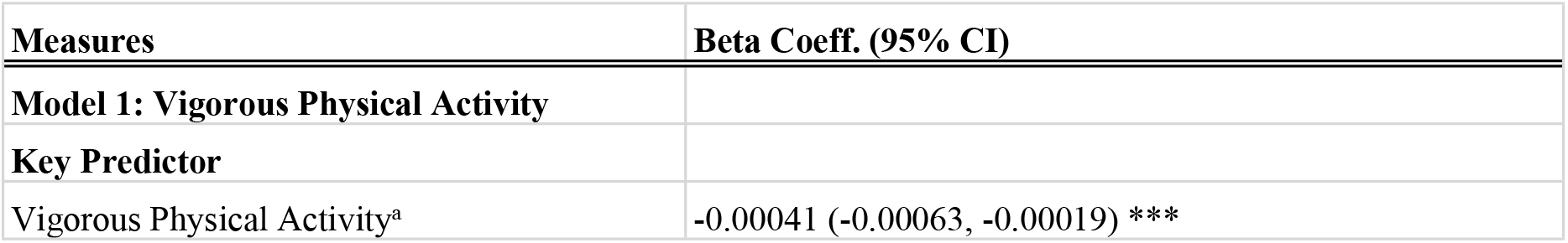
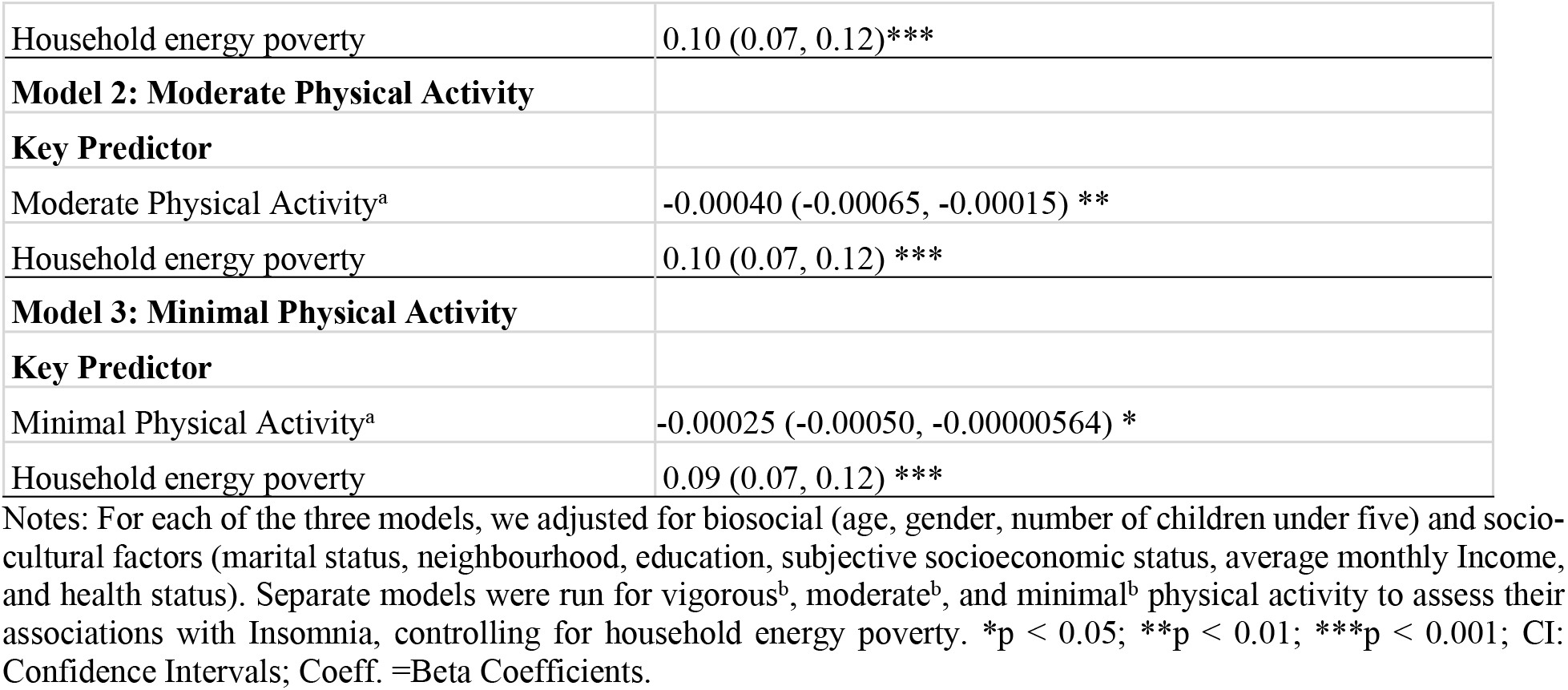
Multivariate regression models assessing the effect of each level of physical activity on Insomnia.

Figure one presents findings that show the main effect of household energy poverty was associated with worse sleep outcomes (β = 0.11, 95% CI: 0.08, 0.14). The main effect of vigorous physical activity showed a decrease in Insomnia (β = -0.00018, 95% CI: -0.00075, 0.00039). The interaction between household energy poverty and vigorous physical activity was associated with lower insomnia scores, although the association was not statistically significant (β = - 0.00001, 95% CI: -0.00004, 0.00001). Similar results were found for the interaction between the other two forms of physical activity (moderate and minimal) and energy poverty. The main effect of energy poverty was consistently robust, while the interaction between energy poverty and physical activity was consistently not significant.

## Discussion

This study aimed to examine the relationship between household energy poverty and insomnia among people living with HIV in Kenya, while accounting for the moderating role of physical activity levels (vigorous, moderate, and minimal). Our findings show that energy poverty worsens Insomnia, but physical activity, regardless of its level, leads to a decline in Insomnia. The moderation effect of all three levels of physical activity was not statistically significant.

A major finding of this study is the significant effect that household energy poverty has on Insomnia, suggesting that as energy poverty increases, Insomnia also increases, which is consistent with the existing literature (62,63). With minimal research focused on PLHIV (26,64), this study is the first to show that energy poverty among PLHIV worsens their sleep health. This finding also corroborates other studies that have found a link between household energy poverty and adverse health outcomes (65,66).

Another significant finding from this study was the relationship between the different levels of physical activity and Insomnia. Vigorous, moderate, and minimal physical activity were shown to slightly reduce the chances of Insomnia, even after accounting for the effects of household energy poverty. These findings underscore the importance of physical activity as a potential mitigating factor in addressing sleep problems among energy-deprived households. These findings are consistent with studies conducted by Kline et al. (2021) and Obeidat et al. (2024), which show that engaging in physical activity improves sleep health.

The interaction model showed that physical activity may slightly reduce the adverse effects of household energy poverty on Insomnia, suggesting that engaging in vigorous or moderate physical activity may reduce Insomnia among people living in energy-deprived households. Vigorous physical activity showed the most significant decrease in insomnia levels, while moderate and minimal showed weaker effects. Although the interaction was not statistically significant, implementing programs and policies that address the causes of household energy poverty may be more effective in improving sleep outcomes. Future studies should investigate these interactions further using longitudinal data to gain a better understanding of how physical activity interacts with household energy poverty.

Socioeconomic factors such as average monthly income and subjective socioeconomic status were linked with a reduction in Insomnia. This aligns with existing literature that suggests that stronger economic standing can buffer the negative impacts of household energy poverty (67,68). This finding is also consistent with the social gradient of health, which suggests that health outcomes improve as wealth increases and worsen as economic status decreases (69,70). Consistently, an increase in higher socioeconomic status and greater monthly Income is associated with lower levels of Insomnia.

Interestingly, the level of education did not have any significant effect on Insomnia. While previous studies have linked higher educational attainment to better health outcomes, this result suggests that in energy-deprived households, education alone may not be sufficient to serve as a protective factor, thereby reflecting deeper barriers that persist regardless of educational background.

Finally, poorer self-rated health was associated with significantly greater Insomnia, emphasizing the relationship between physical health and sleep quality. This may be attributed to the adverse effects of chronic illness or pain, which can contribute to worse sleep outcomes (71). Additionally, individuals in poor health may experience higher levels of stress, which can disrupt their sleep (72). These findings underscore the need for comprehensive health interventions that address both physical and mental health to enhance sleep outcomes.

Despite the significance of our findings, this study is not without limitations. First, social desirability bias may have influenced participants’ responses, as questionnaires are prone to biases such as overreporting or underreporting due to recall errors (73). Second, the study used a cross-sectional survey design, which involved collecting data at a single point in time, limiting our ability to draw causal conclusions (74). Suggestions for future research should consider using a longitudinal study, in which participants are tracked and data are collected over time, to help establish causal pathways (75). Third, measurement errors in predictor variables can impact moderation models (76). Future research should consider using latent variable modelling to reduce the measurement errors.

## Conclusion

This study revealed that household energy poverty is significantly associated with Insomnia among PLHIV in Kenya. Also, it finds that any level of physical activity could reduce Insomnia and improve sleep health. These findings highlight the need for interventions such as the promotion of physical activity in community-based HIV programs as a low-cost behavioural strategy to improve sleep outcomes among vulnerable populations. Concurrently, policy recommendations should include expanding access to clean energy in low-resource settings. Additionally, public health interventions should also focus on reducing socioeconomic disparities, as greater Income and higher perceived social status were associated with reduced Insomnia. In summary, future programs should be holistic, targeting environmental and behavioural risk factors to reduce the burden of Insomnia and improve the quality of life for PLHIV in sub-Saharan Africa.

## Data Availability

The data that support the findings of this study are available on request from the corresponding author. The data are not publicly available due to privacy or ethical restrictions.

N/A

## Acknowledgements

We want to thank the Pamoja Community-Based Organization (CBO) in Kisumu County, Kenya, for their support in the implementation of this study. We are also grateful to the participants from Seme and Kisumu West sub-counties.

